# The *All of Us* Research Program: data quality, utility, and diversity

**DOI:** 10.1101/2020.05.29.20116905

**Authors:** Andrea H. Ramirez, Lina Sulieman, David J. Schlueter, Alese Halvorson, Jun Qian, Francis Ratsimbazafy, Roxana Loperena, Kelsey Mayo, Melissa Basford, Nicole Deflaux, Karthik N. Muthuraman, Karthik Natarajan, Abel Kho, Hua Xu, Consuelo Wilkins, Hoda Anton-Culver, Eric Boerwinkle, Mine Cicek, Cheryl R. Clark, Elizabeth Cohn, Lucila Ohno-Machado, Sheri Schully, Brian K. Ahmedani, Maria Argos, Robert M. Cronin, Christopher O’Donnell, Mona Fouad, David B. Goldstein, Philip Greenland, Scott J. Hebbring, Elizabeth W. Karlson, Parinda Khatri, Bruce Korf, Jordan W. Smoller, Stephen Sodeke, John Wilbanks, Justin Hentges, Christopher Lunt, Stephanie A. Devaney, Kelly Gebo, Joshua C Denny, Robert J. Carroll, David Glazer, Paul A. Harris, George Hripcsak, Anthony Philippakis, Dan M. Roden, On behalf of the *All of Us* Research Program

## Abstract

**Importance:** The *All of Us* Research Program hypothesizes that accruing one million or more diverse participants engaged in a longitudinal research cohort will advance precision medicine and ultimately improve human health. Launched nationally in 2018, to date *All of Us* has recruited more than 345,000 participants. *All of Us* plans to open beta access to researchers in May 2020.

**Objective:** To demonstrate the quality, utility, and diversity of the *All of Us* Research Program’s initial data release and beta launch of the cloud-based analysis platform, the cloud-based Researcher Workbench.

**Evidence:** We analyzed the initial *All of Us* data release, comprising surveys, physical measurements (PM), and electronic health record (EHR) data, to characterize *All of Us* participants including self-reported descriptors of diversity. Data depth, density, and quality were evaluated using medication sequencing analyses for depression and type 2 diabetes. Replication of known oncologic associations with smoking exposure ascertained by EHR and survey data and calculation of population-based atherosclerotic cardiovascular disease risk scores demonstrated the utility of data and platform capability.

**Findings:** The beta launch of the *All of Us* Researcher Workbench contains data on 224,143 participants. Seventy-seven percent of this cohort were identified as Underrepresented in Biomedical Research (UBR) including over forty-eight percent self-reporting non-White race. Medication usage patterns in common diseases depression and type 2 diabetes replicated prior findings previously reported in the literature and showed differences based on race. Oncologic associations with smoking were replicated and effect sizes compared for EHR and survey exposures finding general agreement. A cardiovascular disease score was calculated utilizing multiple data elements curated across sources. The cloud-based architecture built in the Researcher Workbench provided secure access and powerful computational resources at a low cost. All analyses have been made available for replication and reuse by registered researchers.

**Conclusions and Relevance:** The *All of Us* Research Program’s initial release of cohort data contains longitudinal and multidimensional data on diverse participants that replicate known associations. This dataset and the cloud-based Researcher Workbench advance the mission of *All of Us* to make data widely and securely available to researchers to improve human health and advance precision medicine.

## Introduction

The *All of Us* Research Program (*All of Us*) is a longitudinal cohort study aimed at advancing precision medicine and improving human health through enrollment of one million or more diverse individuals across the United States^1^. The National Institutes of Health (NIH) defines precision medicine as the “approach for disease treatment and prevention that takes into account individual variability in genes, environment, and lifestyle for each person”^2^. The conceptual basis for *All of Us*, laid out by the Precision Medicine Initiative Working Group Report to the Advisory Committee to the Director of the National Institutes of Health, centers on recognizing the foundation of biomedical research advanced by prospective cohort studies is accommodation of rapidly expanding data sources including electronic health records (EHRs). This model can also increase the engagement of participants by recognizing study enrollees as partners^3^. This transformation of biomedical research to gather diverse types of data on engaged participants requires novel infrastructure to enable both hypothesis-generating approaches as well as traditional hypothesis testing by various types of researchers, with the ultimate goal of improving individualized care and outcomes. This infrastructure requires secure data storage and facile access to data and tools to test diverse hypotheses.

*All of Us* launched national recruitment in May 2018 and as of May 2020 had enrolled over 345,000 participants, of whom 270,000 have provided biospecimens and survey data. A large multi-disciplinary consortium supports the program, with enrollment centers in varied settings including health provider organizations and community partners. Specific emphasis in the program has been placed on recruiting participants from groups that have been historically underrepresented in biomedical research and at this time over 75% of participants are identified as such^4^. *All of Us* is committed to engaging participants longitudinally ensuring access to their own data and results of research, including support for a participant partnership program to inform the direction of the program and research processes.

Multiple data sources are combined to gather information on *All of Us* participants including survey responses through online health surveys, physical measurements (PM), data from EHRs, and collection of blood or saliva biospecimens. The generation of genomic data is scheduled to start in mid-summer of 2020. A cloud-based Researcher Workbench^5^ has been developed to democratize access for researchers by eliminating requirements for large local infrastructure and to enhance data security by minimizing individual data copies^6^. The platform is designed to meet the FAIR principles of research – Findable, Accessible, Interoperable, and Reusable^7^. Additionally, *All of Us* has developed policies to lower barriers to data access including a passport model granting access based on the identity of the researcher instead of the more common mode of approval based on a single project proposal.

Specific scientific goals and expected timelines have been laid out by *All of Us*, including early, iterative data release and the establishment of demonstration projects showing the quality, usefulness, validity, and diversity of the research data set and platform^8,9^. The data set described here includes survey, measurement, and EHR data on over 220,000 initial participants, now released with the platform two years after the launch of national enrollment. A core value of *All of Us* is to ensure equal access to the data by researchers; therefore, demonstration work was designed to describe the cohort, and replicate previous research findings for validation, and not to make novel discoveries. The first project was designed to include descriptive overviews including defining Underrepresented in Biomedical Research (UBR) populations from survey response data, interrogation of electronic health data showing sequence of medication therapy across common diseases, a phenome-wide examination of the implications of smoking using multiple data sources, and calculation of baseline cardiovascular disease risk scores.

Here, we describe the first demonstration project using the data and tools in the initial release of the *All of Us* Researcher Workbench. All methods, cohorts used, and relevant analytical code have been made available in the Researcher Workbench for replication and reuse by approved researchers.

## Methods

### Protocol

The goals, recruitment methods and sites, and scientific rationale for *All of Us* have been described previously^1^. Participants consent to the study and authorize sharing of the EHRs through an online portal or smartphone application after which they can answer health surveys, share digital health data (such as FitBit and Apple HealthKit), and can view their study information. Through in-person visits, participants can be invited to contribute biospecimens and undergo physical measurements.

The first three surveys available immediately at the time of enrollment included “The Basics” (socio-demographic questions), “Overall Health” (general health overview), and “Lifestyle” (tobacco, alcohol, and drug use related questions)^10^. The second set of three surveys are made available 90 days after enrollment and include “Healthcare Access & Utilization” (how participants access and utilize the healthcare system), “Family History” (the participant’s family health history), and “Personal Medical History” (a participant’s medical history). The details of these surveys are available in the Survey Explorer found in the Research Hub, a website designed to support researchers^11^. Each survey includes branching logic and all questions are optional and may be skipped by the participant. At the enrollment visit, physical measurements (PM) are recorded including systolic and diastolic blood pressure, height, weight, heart rate, waist and hip measurement, wheelchair use, and current pregnancy status.

The third main data type collected prior to the curation of this dataset release was structured EHR data. This data type is transferred from enrolling sites to the Data and Research Center at least once per quarter. Additionally, the program has launched capabilities for Fitbit data sharing and participant-mediated EHR transfer via Apple HealthKit integration; these data were not acquired prior to the curation of the dataset used for this study and will be included in future releases.

### Data curation and privacy methodology

All three data types (surveys, PM, and EHR) are mapped to the Observational Medical Outcomes Partnership (OMOP)^12^ common data model v 5.2 maintained by the Observational Health Data Sciences and Informatics (OHDSI) collaborative^13^. Where the model did not support necessary concepts, custom concepts were added in collaboration with the OHDSI community, were linked to existing concepts where possible, and are documented in the open source Athena resource, a repository of vocabularies used in OMOP and supported by Odysseus Data Services^14^. Participants were included in the beta launch curated data repository (CDR) if they responded to at least the first survey, “The Basics.”

To protect participant privacy, a series of data transformations are applied including data suppression of codes with a high risk of identification such as military status, generalization of categories including age, sex at birth, gender identity, sexual orientation, and race, and date shifting by a random number of days from 1 to 365 implemented consistently across each participant record. General conformance rules were applied to meet the standard conventions of the OMOP data model including dropping invalid dates and extreme values resulting in a base version of the CDR. Additional cleaning steps for selected lab data (from EHRs) and PMs were performed to standardize units and values resulting in the processed CDR, on which the analyses presented here were performed. Documentation on privacy implementation and creation of the CDR is available in the Research Hub at http://www.researchallofus.org and available in the *All of Us* Registered Tier CDR Data Dictionary^15^.

### Platform

The dataset was accessed through the *All of Us* Researcher Workbench platform, a cloud-based analytic platform custom built by the program for approved researchers. The Workbench is built on top of the Terra platform^16^, which is also utilized for a number of other NIH-funded studies including the NCI Cloud Resources, the NHLBI BioData Catalyst, and the NHGRI AnVIL^17–19^. The Workbench exceeds Federal Information Security Management Act (FISMA) Moderate security standards and undergoes routine security testing^20^. The *All of Us* Researcher Workbench platform includes Workspaces that are project-specific spaces featuring a description of the project and permitting sharing among teams of collaborators. Workspaces include access to graphical interface tools to select participants (a “cohort builder”) and data elements for storage and analysis in Jupyter Notebooks^21^. The notebooks currently enable use of saved datasets and direct query using R and Python 3 programming languages. Access to the Researcher Workbench and data are free. Compute and storage accrue usage cost. The Researcher Workbench uses Google Compute Engine for computational resources in the cloud and Google Cloud Storage for storage in the cloud.

### Access

All researchers who accessed the data for analyses were authorized and approved via a 3-step process that included registration, completion of ethics training, and attestation to a data use agreement^5^. Approval to use the dataset for the specified demonstration projects was obtained from the *All of Us* Institutional Review Board. Results reported are in compliance with the *All of Us* Data and Statistics Dissemination Policy disallowing disclosure of group counts under 20 to protect participant privacy^22^.

### Descriptive visualizations

Age displayed reflects age when the CDR version used in this report was generated in the fall of 2019. Presence of a data type survey, PM, or EHR was counted if at least one observation is present within each category. Regarding race and ethnicity, participants were asked “Which categories describe you? Select all that apply. Note, you may select more than one group”, in the “The Basics” survey. Responses were mapped to the race variable in the OMOP Person table directly for the responses ‘White’, ‘Asian’, and ‘Black, African American, or African’, and responses ‘Middle Eastern or North African’, and ‘Native Hawaiian or other Pacific Islander’ were generalized to ‘Other single population’. Currently, all participants responding ‘American Indian or Alaska Native’ have been removed from the CDR as *All of Us* goes through official Consultation with tribal leaders on the research use of data. Participants choosing any two of the categories were labeled ‘More than one population’. Skipped questions were omitted, and the responses ‘None of these fully describe me’ and ‘I prefer not to answer’, or nonresponse to these categories were individually mapped in the data model and grouped as ‘Not specified’ for visualization for the analyses presented here. The ‘Not specified’ group includes participants who chose ‘Hispanic, Latino, or Spanish’. This response was mapped to the ethnicity variable, allowing reflection of both race and Hispanic ethnicity. Program designations of status as UBR were adapted to data available in the CDR^23^.

### Treatment pathway visualization

The order of treatment prescribed after common disease diagnosis was determined for type 2 diabetes (T2D) and depression to demonstrate medication mapping and use of hierarchies in the OMOP common data model. This work replicates analyses published by the OHDSI consortium on a larger dataset^24^. For each condition, the time of earliest diagnosis was identified and medications were extracted using the OMOP hierarchy as in the previously published work. Medications were then grouped into generalized classes based on their main ingredient using the Anatomical Therapeutic Chemical (ATC) Classification^25^. Participants were included if their first medication related to the disease was prescribed after the earliest diagnosis code for that disease, they had two or more diagnosis codes for the disease, and they had at least three years of medication records with at least a single structured occurrence of the drug. We determined the number of participants whose monotherapy was the most common first medication in any given year between 2000 to 2016. Each of these analyses was performed separately on the participants identified as White race and compared to those included in the UBR category with any non-White response. A chi-square test was used to compare medication sequences between races.

### Phenome-wide association with smoking study

A cancer phenome-wide association study (PheWAS) was performed to replicate known associations with smoking and compare effect sizes of smoking exposure gathered from EHR billing codes with smoking exposure determined from survey data^26,27^. To define ever smoking exposure from EHR data (‘EHR Ever Smoker’), we identified all participants with at least two instances on separate calendar days of ICD9CM codes 305.1* (Tobacco use disorder), 649.0* (Tobacco use disorder complicating pregnancy, childbirth, or the puerperium), V15.82 (History of Tobacco Use), and 989.84 (Toxic effect of tobacco), or ICD10CM codes Z72.0 (Tobacco Use), Z71.6 (Tobacco abuse counseling), O99.33* (Tobacco use disorder complicating pregnancy, childbirth, and the puerperium), F17.2* (Nicotine Dependence) excluding F17.22* (Nicotine Dependence, chewing tobacco), and T65.2* (Toxic effect of tobacco and nicotine) excluding T65.21* (Toxic effect of chewing tobacco). To define never smokers from EHR data (‘EHR Never Smoker’), zero occurrences of the defining codes above and at least one other ICD-9-CM or ICD-10-CM code that was not T65.21 or T65.2 was required. To define smoking exposure from survey data (‘Survey Ever Smoker’), the “Lifestyle” survey responses were used. Specifically, the response to “Have you smoked at least 100 cigarettes in your entire life” was used to include participants as a Survey Ever Smoker, and the branching logic question “Do you now smoke cigarettes every day, some days, or not at all?” with the response “Every day” to designate current smokers. Conversely, participants answering “No” to the 100 cigarettes question were included as ‘Survey Never Smokers’, and participants skipping the question were excluded. The logistic regression model used in the PheWAS analyses was implemented using the statsmodels Python module, optimized for compute efficiency, and made flexible for reuse with variable inputs. The analysis was corrected for age at last code occurrence, sex at birth, race and ethnicity as generalized from survey responses, EHR length as reflected by time between first and last billing code, and unique billing codes per record.

To compare the phenome-wide associated effects found in the respective PheWAS analyses to prior results, we searched PubMed for meta-analyses that produced effects comparable to the odds ratios produced by the logistic regressions. We first searched PubMed for all meta-analyses related to smoking using the R package easyPubMed and query “(tobacco[TI] OR smoking[TI]) AND meta-analysis[All Fields]”), finding 1686 results. Then computable restrictions were applied including limitation to active smoking-exposed individuals and excluding genetic and smoking cessation studies, resulting in 491 studies. Manual review then included only those titles with phenotypes represented in the PheCode ontology resulting in 265 papers covering 133 unique phenotypes^28–30^. We then restricted to only those meta analyses that could be matched with a phenome-wide significant result from at least one of the PheWAS analyses and a comparable effect, finding 75 meta-analyses across 55 unique phenotypes, of which 6 had a phenome-wide significant result that was related to an oncologic outcome with ever smoking exposure. Results were plotted to compare with *All of Us* results for EHR and Survey Ever Smoking phenotypes.

### Cardiovascular disease risk calculation

Ten-year atherosclerotic cardiovascular disease (ASCVD) risk was calculated according to the 2013 Pooled Cohort Equations^31^. Participants were included if aged 40–79 when EHR data were available including total cholesterol (TC), high-density lipoprotein cholesterol (HDL), systolic blood pressure (SBP) and treatment status, diabetes status, and no evidence of existing ASCVD. The codes used to identify ASCVD outcome, diabetes, hypertension, and treatment are available in Supplemental Table 1. We removed values outside the valid ranges for the score: TC 130–320, HDL 20–100, and SBP 90–200 mm Hg. Measurements were included within one year of the most frequently available variable, SBP. If multiple measures were in the window, the median was used. Current smoking status was taken from the participants’ survey response within “Lifestyle” branching logic to age started smoking and age stopped smoking to determine if smoking occurred during the score calculation. The beta coefficients for African American race were used if the participant selected only “Black, African American, or African”, and White if selected only “White” in “The Basics” survey. Any other response, multiple responses, or skip was designated as ‘Other’ here which also uses the White race beta coefficients in the ASCVD model. To compare risk scores among race groups, the Kruskal-Wallis H and Mann-Whitney U non-parametric tests were used. To calculate the optimal risk, we used 170 for total cholesterol, 50 for HDL, 110 for SBP, and status as non-smoker, non-hypertensive, and non-diabetic.

## Results

### Demographics of the beta-launch data set

The beta launch of the Researcher Workbench includes de-identified data from 224,143 total participants. Figure 1A displays an overview of the data types available, separating the surveys into Part 1 which includes the first three surveys participants completed, and Part 2 which includes the second set of three surveys that were made available 90 days after enrollment. Note that because *All of Us* de-identification requires a random date shift within the past year, an artificial leveling off appears and survey data is represented before the start of program enrollment. By design all 224,143 participants have data from “The Basics” survey. The second two surveys, “Overall Health” and “Lifestyle” have 218,209 and 216,799 participant responses, respectively. The final three surveys distributed after enrollment, “Healthcare Access & Utilization”, “Family History”, and “Personal Medical History”, have 45,676, 41,468, and 39,200 participant responses, respectively. Of those with any survey response, 188,549 have at least one PM recorded, and 127,133 have any EHR data included. The total number of participants who have any survey response, PM, and EHR data is 126,457. Additional breakdowns of individual data types are shown in Supplemental Figure 1. Figure 1B shows the historical availability of EHRs, including a breakdown of structured domains within the OMOP CDM.

**Figure 1:**
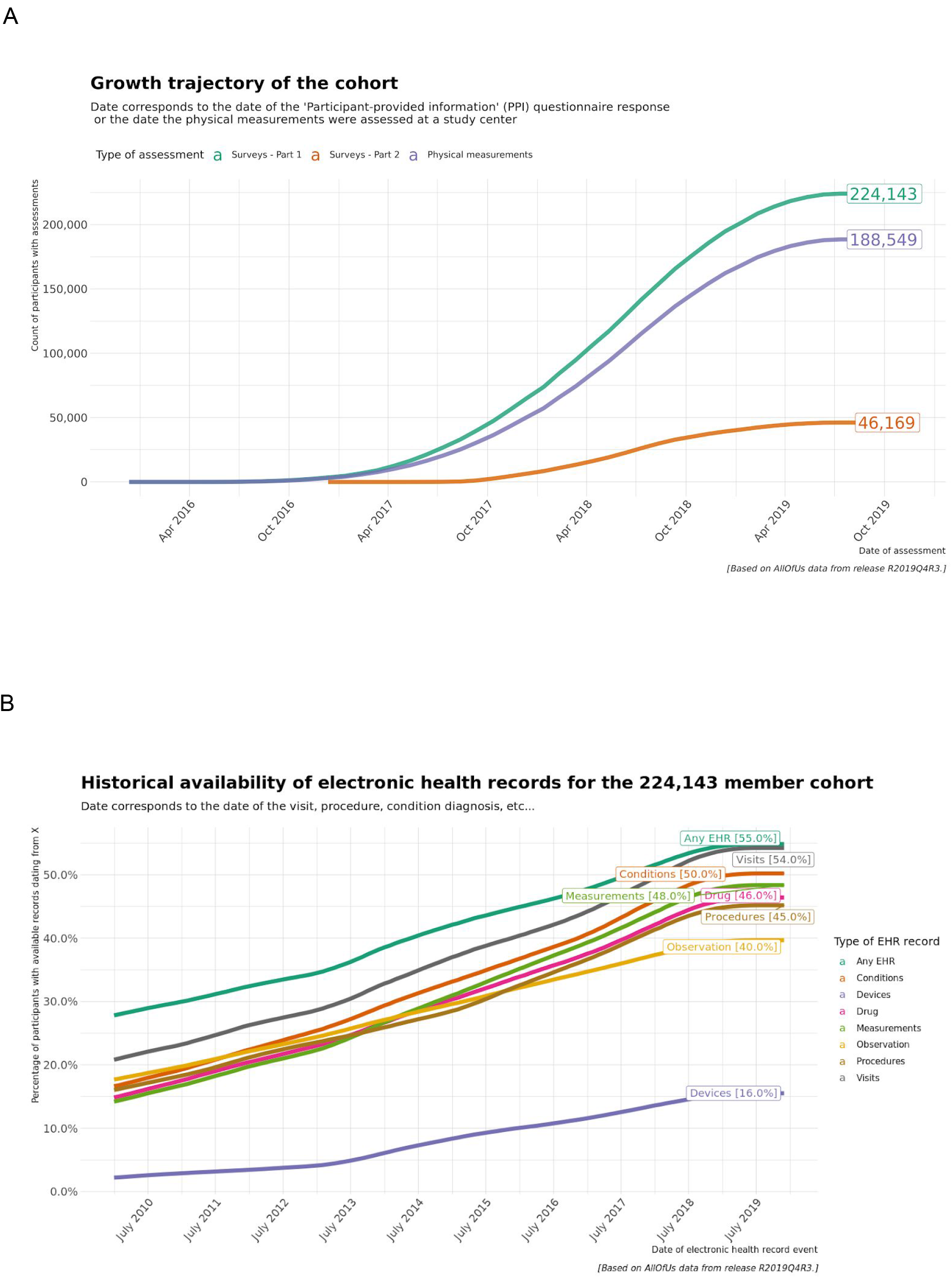
Overview of data types included in the beta release CDR. A: Growth trajectory of participant data types after enrollment. Survey Part 1 (green) includes “The Basics”, “Lifestyle”, and “Overall Health” surveys; Survey Part 2 (red) includes “Personal Medical History”, “Health Care Access and Utilization”, and “Family Medical History; Physical measurement accrual is shown in purple. Note the flattening is artificial due to the random date shift introduced to protect participant privacy. B: Historical availability of participants’ electronic health records (EHR). The green line reflects any EHR record, and the various colors represent domains within the EHR.

Demographic information included in the dataset is extracted from “The Basics” survey response. Figure 2 shows responses to the question “Which categories describe you? Select all that apply”. Figure 2A describes participant responses as generalized into the permitted race categories. For example, inclusion in ‘White’ indicates no other answers in the race categories were selected. If more than one race was selected, such as ‘White’ and ‘Asian’, the participant was included in ‘More than one population’. The response ‘Hispanic’ was mapped separately to the ethnicity variable. Figure 2B shows that most participants that did not select an answer mapped as race, did select the ‘Hispanic’ response. This mapping allows use of both variables to reflect, for example, ‘Black’ and ‘Hispanic’ while other multiple responses are generalized in ‘More than one population’. Figure 2C shows the distribution of ages within each race, including a higher percentage of older participants in the ‘White’ and ‘Black’ only race. Figure 2D shows the percentage of participants by race and sex at birth demonstrating consistent higher enrollment of women in each group. Figure 3 details additional survey responses into the program definitions of participant status as UBR. Notably over 47 percent of participants identified with a population other than ‘White’ alone and 13 percent of participants identified with a sexual or gender minority group. Overall 77% of participants were included in at least one UBR category.

**Figure 2:**
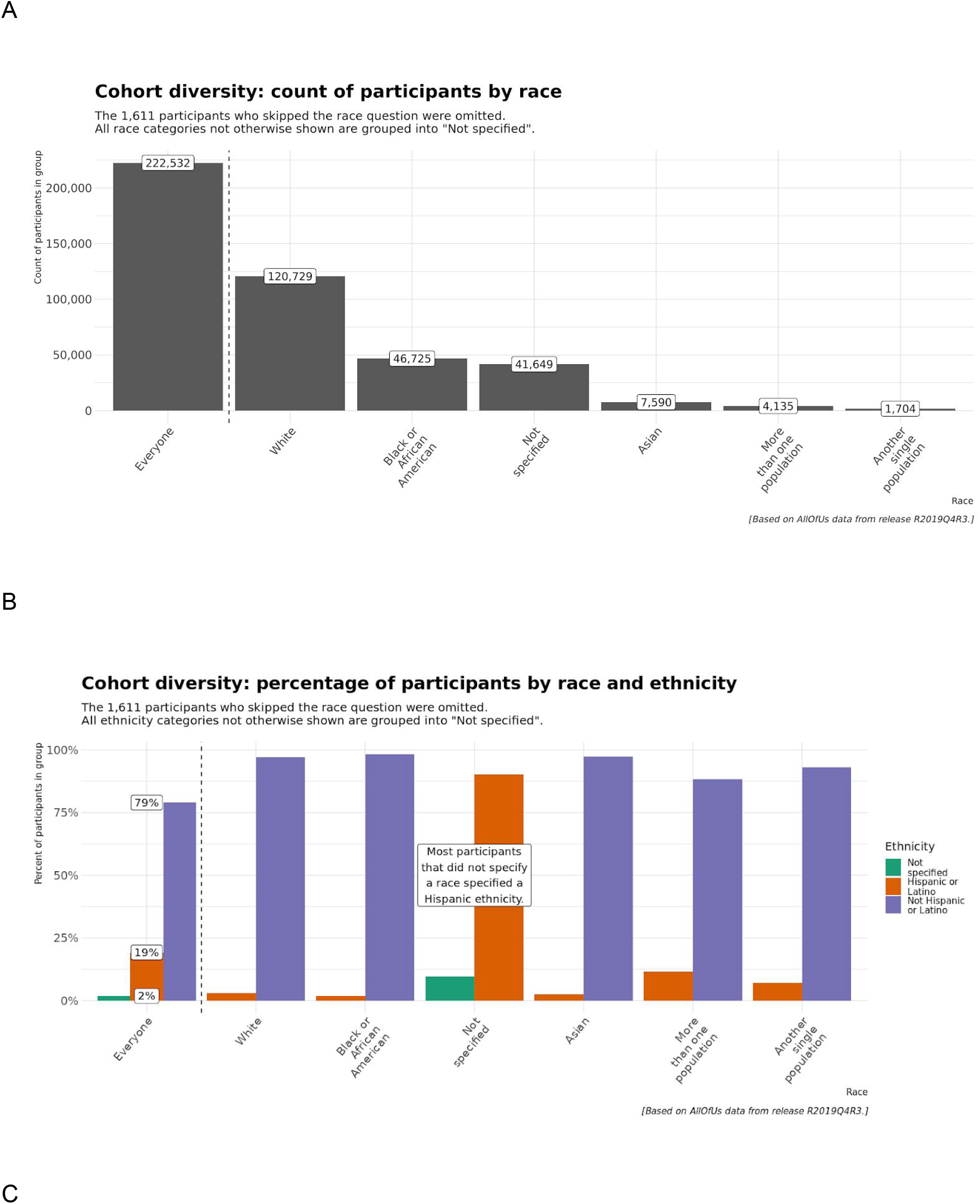
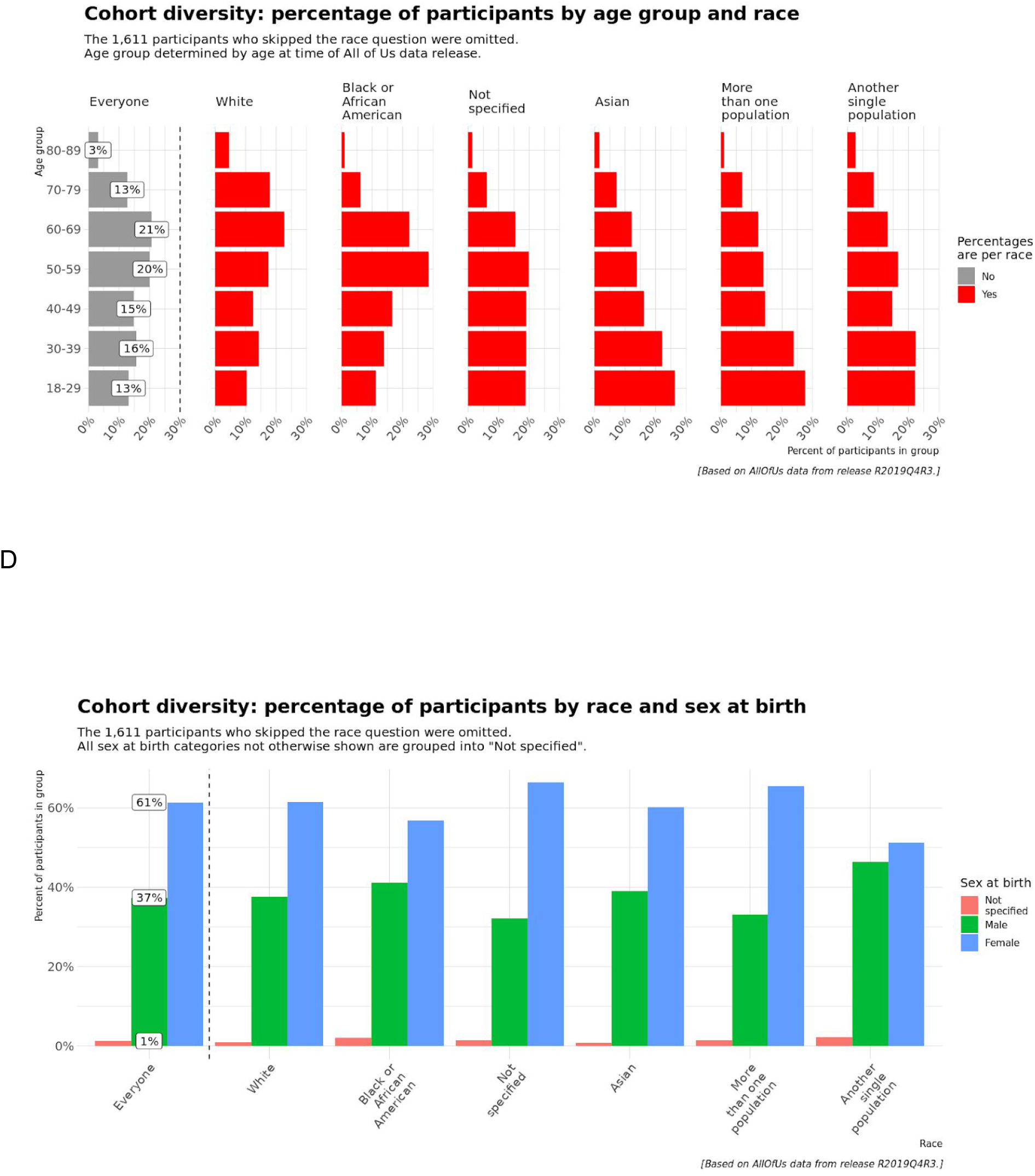
Participant age, race, and ethnicity characteristics. A: Count of participants by race options available in “The Basics” including White, African American or Black, Not Specified, Asian, More than one population, and Another single population. B: Percent of participants by race and ethnicity; ethnicity categories available in “The Basics” survey include “Hispanic or Latino” and “Not Hispanic or Latino”. All ethnicity categories not otherwise shown are grouped as “Not Specified”. C: Percent of participants by age group and race; for ages ranging from 18–89 years of age and race options available in “The Basics” survey D: Percent of participants by race and sex at birth; sex at birth categories available in “The Basics” survey include “Male”, “Female” and “Not specified” when category not otherwise specified.

**Figure 3:**
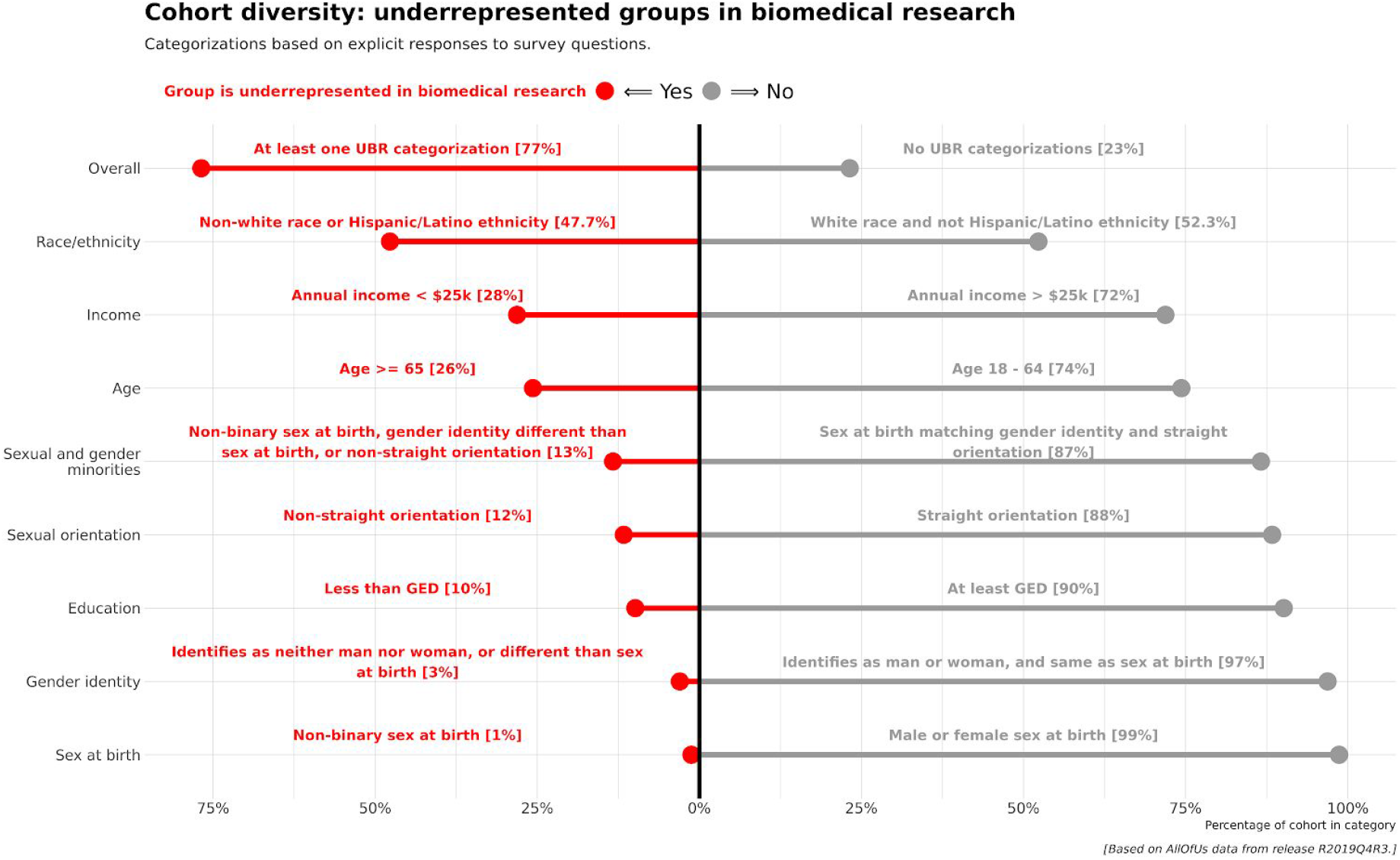
UBR metrics. Depiction of the proportion of participants that are underrepresented in biomedical research (UBR) based on program definitions. A participant is included in the ‘Overall’ category if they meet at least one criterion among the Race/ethnicity, Income, Age, Sexual Orientation, Education, Gender Identity, and Sex at Birth designations. The ‘Sexual and gender minorities’ category shown aggregates any participant with a UBR response to questions on Sexual Orientation OR Gender Identity OR Sex at Birth. GED: General Education Development (i.e., High School Diploma or equivalent)

### Treatment pathway visualization

The numbers of participants meeting inclusion criteria to map treatment pathways were 9,456 total for T2D and 15,776 for depression. The number of participants contributed by individual consortium EHR sites are shown in Supplemental Table 2. The treatment pathway visualizations are shown in Figure 4A-F independently with counts for White and UBR participants. The innermost circle represents the first medication class prescribed, and the circles expanding outward are the second and third medication classes occurring in the EHR after diagnosis. The most common first medication classes were biguanides for T2D and selective serotonin reuptake inhibitors for depression in both White and UBR participants, which match the original OHDSI analysis across much larger sites. The first treatment prescribed for both T2D and depression differed between White and non-White participants (p< 0.001).

**Figure 4:**
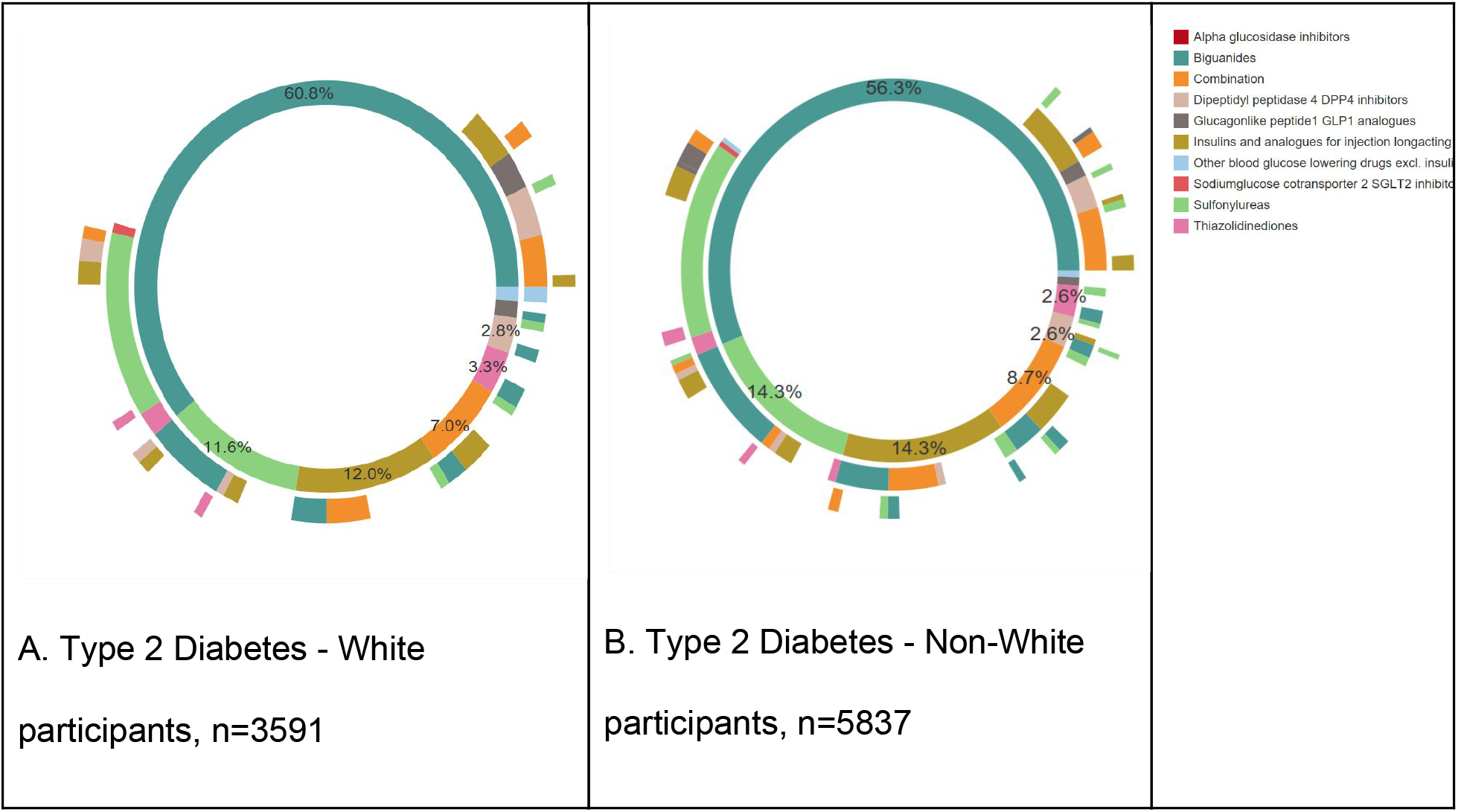
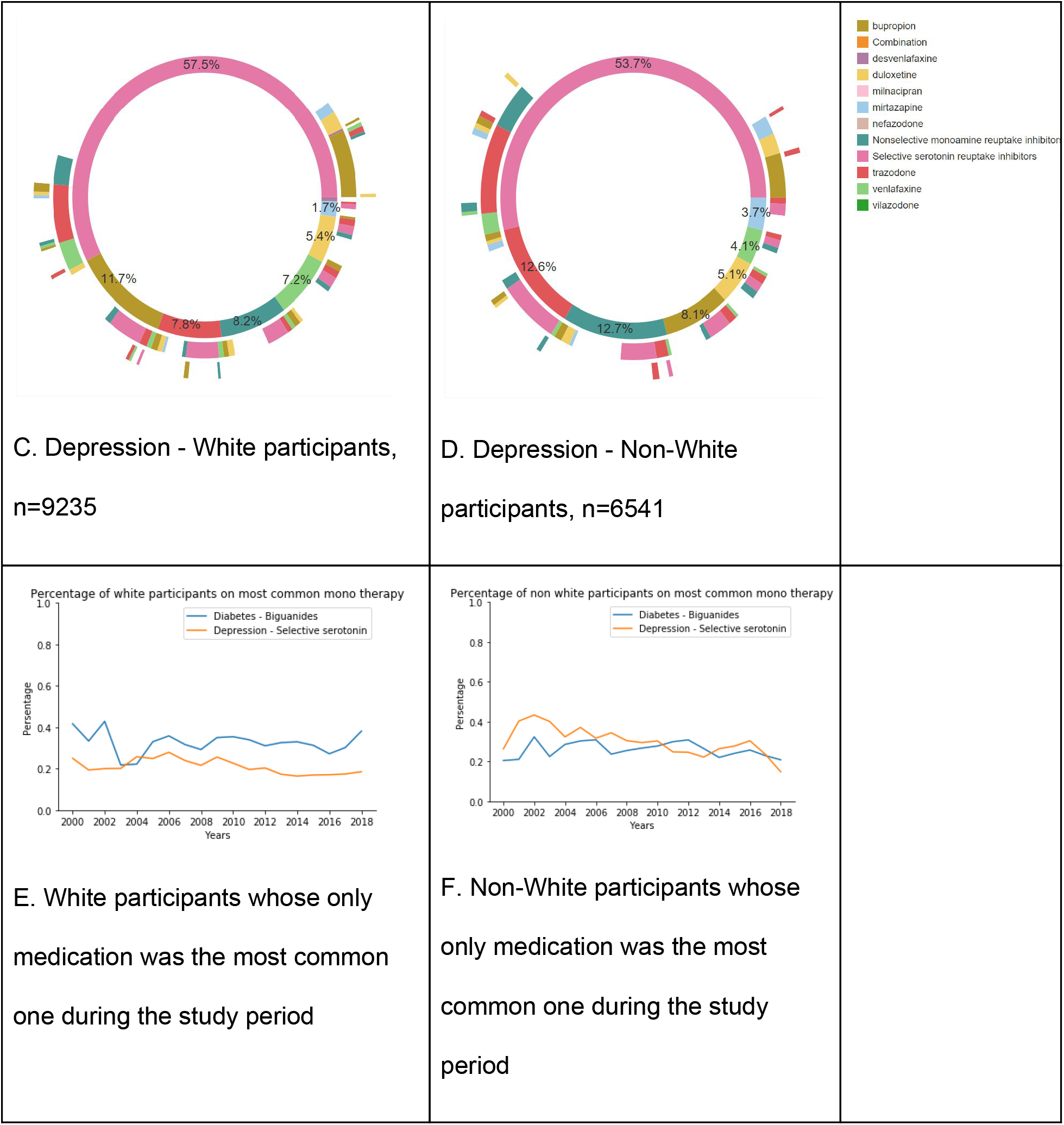
Treatment pathways for T2D and Depression. Medication sequencing for participants who have diabetes and depression grouped by race. A: Anti-diabetic medication sequences for White participants. B: Anti-diabetic medication sequences for non-White participants.C: Antidepressant medication sequences for White participants. D: Antidepressant medication sequences for non-White participants. E: Percentage of White participants who were prescribed one medication that is the most common one from year 2000 to 2018. F: Percentage of non-White participants who were prescribed one medication that is the most common one from year 2000 to 2018. The difference in counts of first antidiabetic in Figures 4A and 4B, and the counts of first antidepressants in Figure 4C and Figure 4D for each medication between white and non-white participants was significant (p-value of chi-square was < 0.05).

### Phenome-wide association with smoking study

We identified 18,335 participants as EHR Ever Smokers and 86,231 EHR Never Smokers using the methods defined above. From the survey responses regarding smoking, 85,929 participants were identified as Ever Smokers and 38,906 participants as Current Smokers. The overlap of these participants is shown in Supplemental Table 3. The PheWAS results for EHR Ever Smoking and Survey Ever Smoking, are shown in Figure 5a and 5b, with each point sized proportionally to their corresponding effect size. Table 1 provides results of the top five EHR non-protective and protective phenome-wide significant effects matched to their phenome-wide significant result from the Survey Ever smoking PheWAS. An expanded results list for each is included in Supplemental Table 4. The top phenotypes for which ever smoking was not protective include respiratory cancers and bladder cancer, both known associations. Alternatively, we see that smoking was protective for cutaneous-related neoplastic outcomes, which has also been reported in the literature^32–34^. A comparison between the effect sizes seen in EHR results versus Survey Ever Smokers and Survey Current Smokers is shown in Figure 5c. The comparison of effect sizes seen in *All of Us* data to literature studies is shown in Figure 5d. The phenotypes shown are sorted by the natural logarithm of effect size reported in the comparator meta-analysis (either OR, RR, or HR) along with corresponding confidence bounds. Effect sizes and confidence bounds from each PheWAS regression result are shown matched to their corresponding meta-analysis^35–40^). Each phenotype represented has at least one PheWAS result from either survey or EHR that is concordant in the direction of the overall effect size. Also, in 5 out of 6 of the phenotypes, there is at least one PheWAS result whose confidence interval overlaps with the confidence interval reported in the corresponding meta-analysis.

**Table 1.** PheWAS effect sizes.

| PheCode | Description | EHR Ever Smoking<br>OR (95% CI) | Survey Ever<br>Smoking<br>OR (95% CI) |
| --- | --- | --- | --- |
| Top 5 Increased risk effects |  |  |  |
| 165.10 | Cancer of the bronchus; lung | 4.94 (4.11, 5.95) | 3.19 (2.65, 3.84) |
| 165.00 | Cancer within the respiratory system | 4.94 (4.12, 5.92) | 3.15 (2.62, 3.78) |
| 189.21 | Malignant neoplasm of bladder | 2.36 (1.87, 2.98) | 1.76 (1.42, 2.18) |
| 149.00 | Cancer of the larynx, pharynx, nasal cavities | 2.26 (1.71, 2.97) | NS |
| 189.20 | Cancer of bladder | 2.25 (1.8, 2.82) | 1.69 (1.37, 2.08) |
| Top 5 Decreased risk effects |  |  |  |
| 217.0 | Vascular hamartomas and non-neoplastic nevi | 0.51 (0.42, 0.62) | 0.55 (0.48, 0.64) |
| 217.1 | Nevus, non-neoplastic | 0.52 (0.43, 0.64) | 0.57 (0.49, 0.66) |
| 216.0 | Benign neoplasm of skin | 0.53 (0.49, 0.58) | 0.62 (0.58, 0.66) |
| 228.1 | Hemangioma of skin and subcutaneous tissue | 0.57 (0.47, 0.68) | 0.65 (0.57, 0.74) |
| 228.0 | Hemangioma and lymphangioma, any site | 0.71 (0.62, 0.81) | 0.69 (0.62, 0.77) |

**Figure 5:**
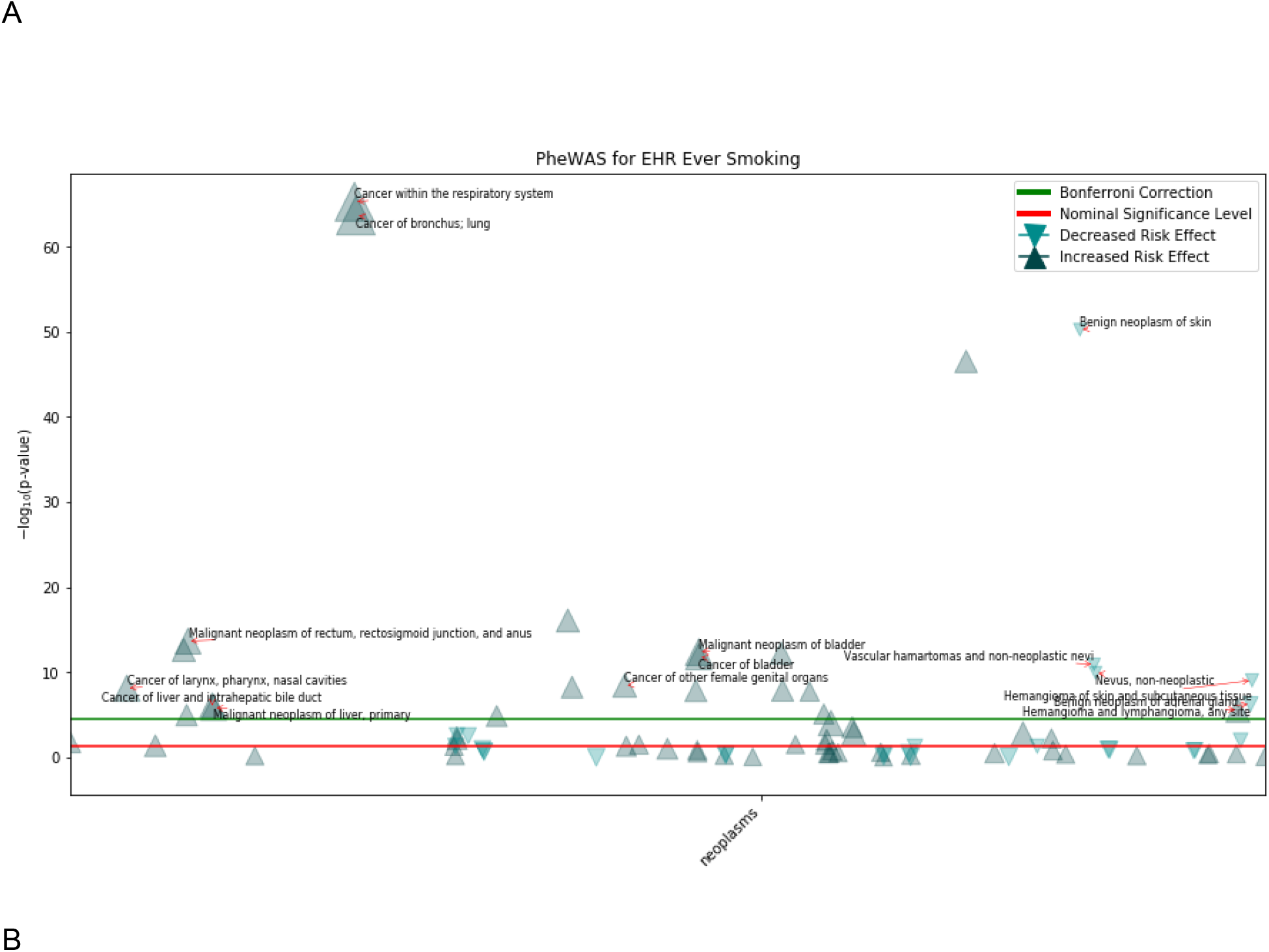
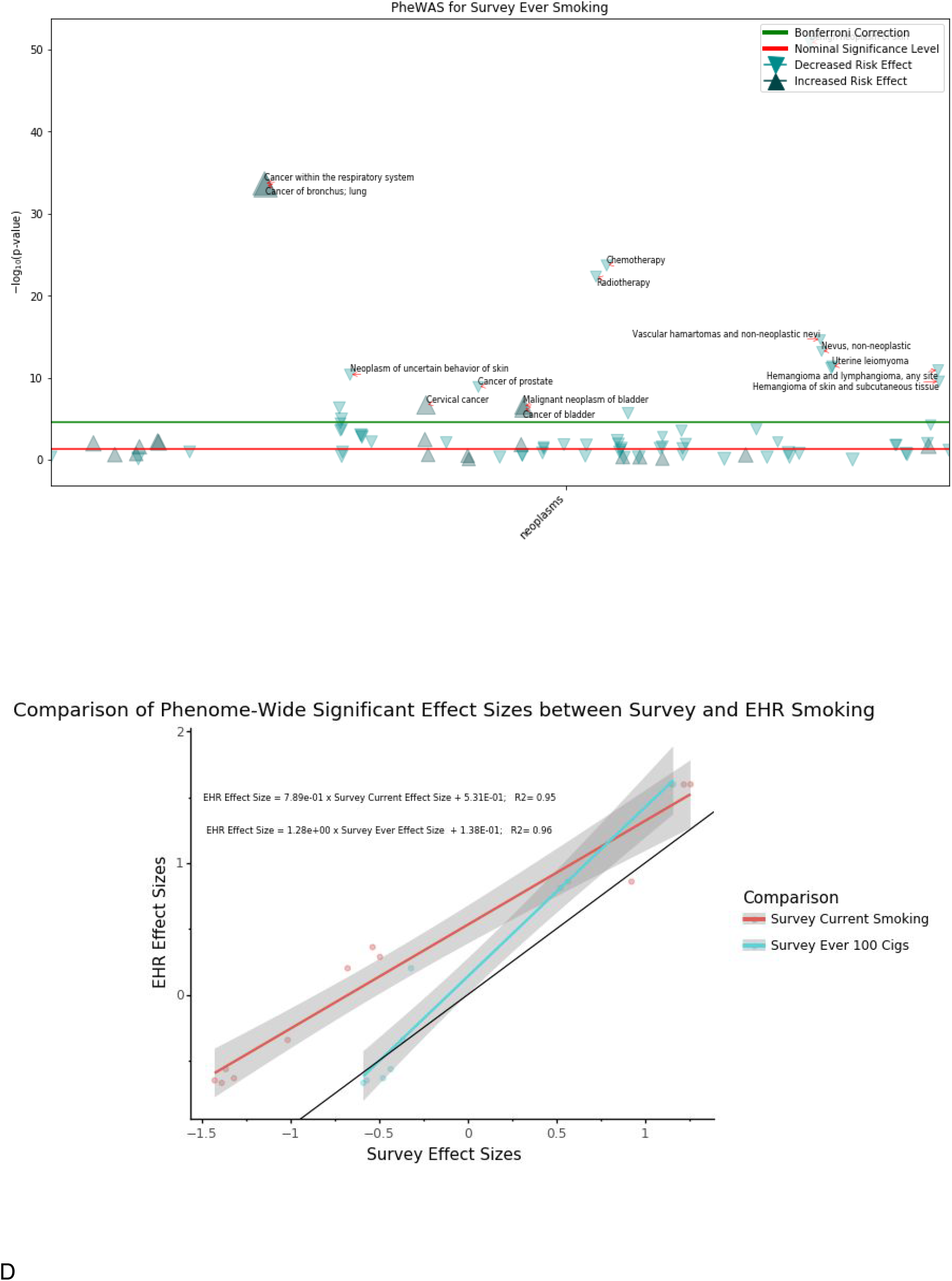
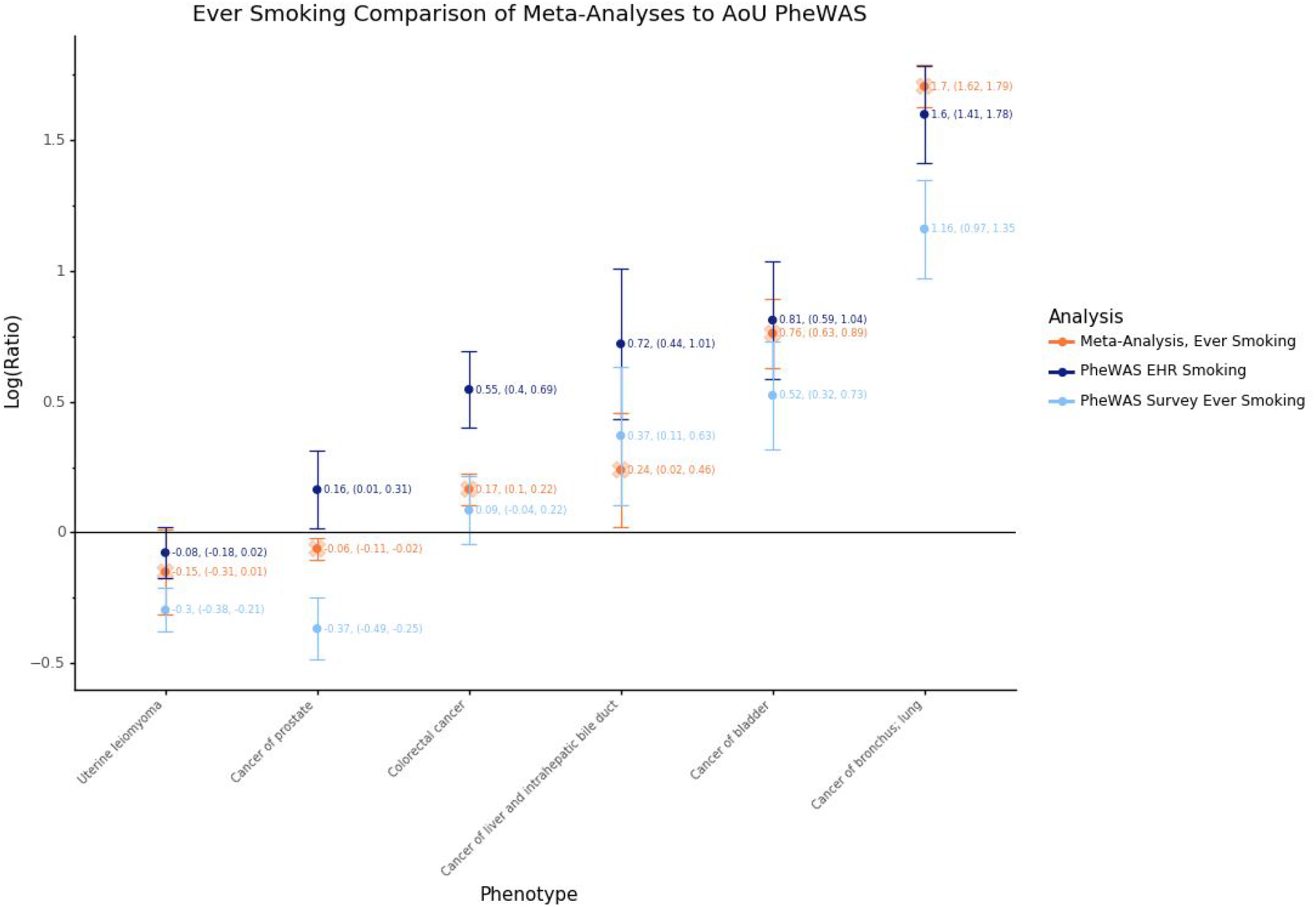
PheWAS Ever Smoking EHR and Survey comparison. A: Manhattan plot for Oncologic PheWAS using EHR Ever-Smoking as exposure. Location of triangles corresponds to phecode numeric value and –log10(p-value) of correspond to top protective and non-protective phenome-wide significant results. B: Manhattan plot for oncologic the corresponding logistic regression. Sizes of data points are proportional to the ever smoking effect size. Labels correspond to top protective and non-protective phenome-wide significant results. B: Manhattan plot for oncologic PheWAS using Survey Ever-Smoking as exposure. Location of triangles corresponds to phecode numeric value and −log10(p-value) of corresponding logistic regression. Sizes of data points are proportional to the ever smoking effect size. Labels correspond to top protective and non-protective phenome-wide significant results. C: Comparison of phenome-wide significant Survey oncologic regression effect sizes (Survey Current Smoking in red and Survey Ever Smoking in blue) compared to EHR phenome-wide significant oncologic effect sizes. Linear regression point estimates are given along with r^2 values. D: PheWAS Ever Smoking EHR (dark blue) and Survey ever smoking (light blue) effect sizes and confidence intervals compared with published meta-analyses (orange). Estimates are presented on the natural log ratio scale (odds ratio (OR), risk ratio (RR), or hazard ratio (HR)). Estimates below the horizontal line represent protective effects and estimates above the line represent non-protective effects.

### Cardiovascular disease risk calculation

There were 25,989 participants with necessary covariates for calculation of the ASCVD risk score prior to observation of any cardiovascular event. Among those participants, 17,340 (66.7%) were female, 5,487 (21.1%) African American, 4,768 (18.3%) other single or two or more races, 15,743 (60.5%) were White, and 9,908 (38.1%) were smokers. Participants with calculated scores average 55.7 (SD ±9.7) years old, and their SBP on average was 127.4 (SD ±14.0). Of these, 4,663 (17.9%) participants had the onset of new CVD within 10 years of measurement (Table 1). Restricting calculation to within one year of enrollment included 6,221 participants, with similar age, race, and smoking distributions, as detailed in Supplemental Table 5. The distribution of risk scores by race is shown in Figure 6. Table 2 shows the score components and composite score broken down by race. Across all three racial groups, scores were significantly different (p< 0.001). In pairwise comparison, risk scores for White and other race participants were lower than African American race (p< 0.001) in both scores calculated at any time and those calculated within a year of enrollment. Other race participant scores were significantly lower than White participants in the scores at any time (p< 0.001), but not in the scores within a year of enrollment (p>0.05).

**Figure 6:**
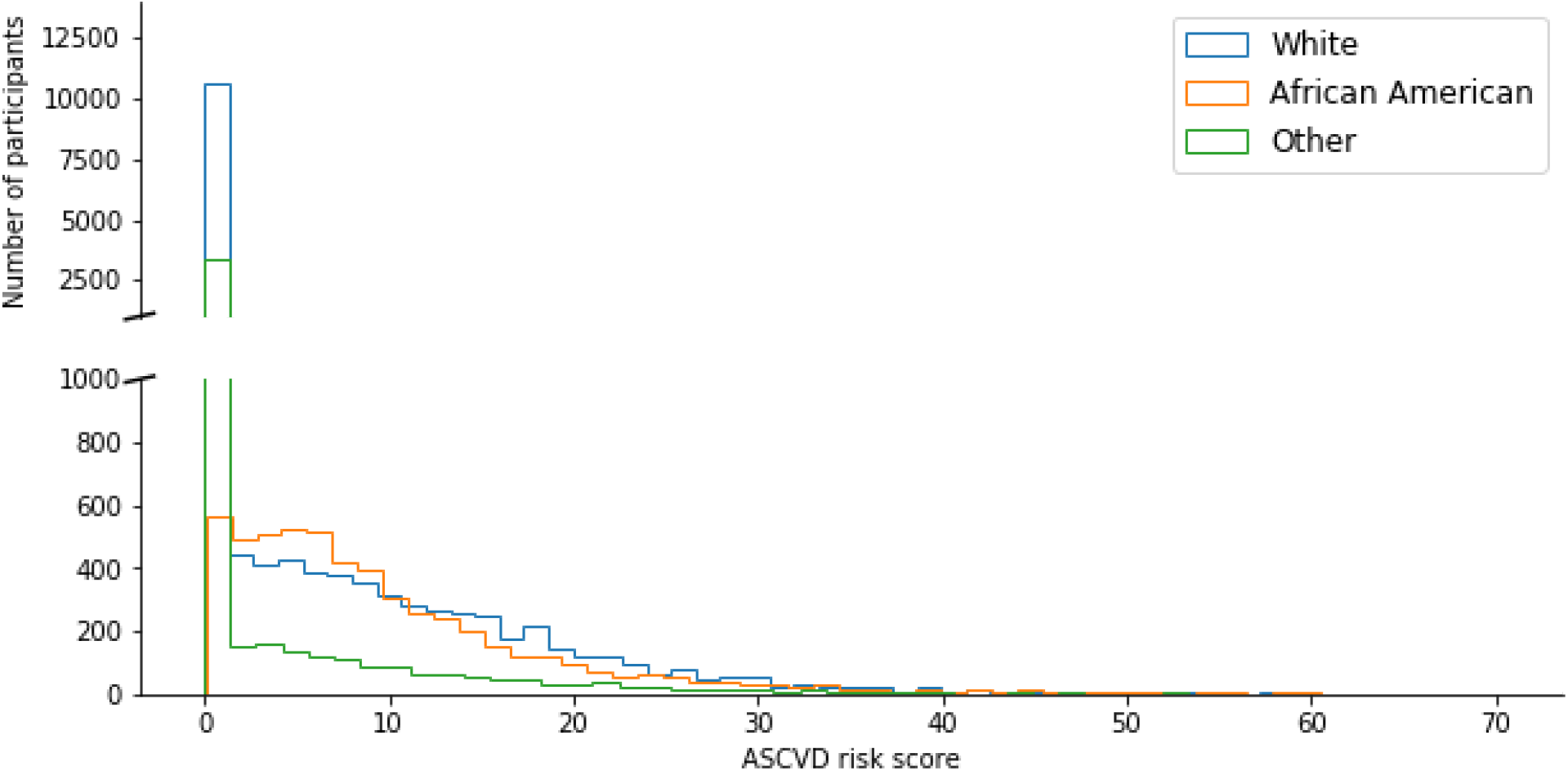
Baseline cardiovascular disease risk calculations. A histogram of the cardiovascular disease risk score for participants with necessary measurements grouped by race group into White, African American, and Other. The difference between the cardiovascular risk scores between the three race groups was statistically significant (p-value for the Kruskal-Wallis H test was 0). Mann-Whitney p-value was < 0.001 when comparing the risk scores for White versus African American participants, Other versus African American, and White versus Other.

**Table 2.** ASCVD scores by race. Descriptive statistics of risk factors used to calculate the ASCVD score for each race group for all participants at any time and those participants whose measurements were available within a year of enrollment in *All of Us*. SD: standard deviation, AF: African American.

|  | All ASCVD |  |  | Scores within a year of AoU enrolment |  |  |
| --- | --- | --- | --- | --- | --- | --- |
|  | Mean (SD), Median |  |  | Mean (SD), Median |  |  |
|  | White | AF | Other | White | AF | Other |
|  | n=15,743 | n=5,487 | n=4,768 | n=3,342 | n=1,574 | n=1,305 |
| Age | 56.86 (9.87),<br>57 | 53.83(8.76),<br>53 | 53.86(9.39),<br>53 | 57.78 (10.47),<br>58 | 55.59 (9.25),<br>56 | 54.28 (9.44)<br>54 |
| Total cholesterol | 191.45(32.14),<br>189 | 185.66(33.03),<br>181 | 188.85(34.424),<br>185 | 191.06<br>(35.59),188 | 187.36<br>(35.29),182 | 189.11(36.74),<br>185 |
| HDL | 55.2(15.55),<br>53 | 53.18(14.55),<br>51 | 50.68 (13.91),<br>49 | 55.1(16.17),<br>53 | 54.44(14.85),<br>52 | 50.92(14.96),<br>49 |
| Systolic blood pressure | 125.70(12.77),<br>125 | 131.69(15.14),<br>130.25 | 127.79(15.42),<br>126.5 | 126.78(16.29),<br>125 | 131.44(17.97),<br>130 | 127.74(17.85),<br>125 |
| Risk | 4.02(7.75), 0 | 9.89 (9.11),<br>7.4 | 3.56(7.65), 0 | 4.67(8.91), 0 | 12.53(11.45),<br>9.05 | 4.22(8.44), 0 |
| Optimal Risk | 2.08 (4.15),0 | 3.15(2.54),<br>2.54 | 1.51(3.42), 0 | 2.33(4.63), 0 | 3.6(2.81), 3.06 | 1.67(3.47), 0 |

### Cost and sharing of analytic methods and code

The total compute execution cost for all analyses described was approximately $96. Supplemental Table 6 indicates the rates and cost of individual analyses. All analyses presented have been made available within the Featured Workspaces section of the Workbench that permits researchers to view and duplicate code for replication of analyses or adaptation for their own use.

## Discussion

A core principle of the *All of Us* program is providing data to researchers quickly, in a manner that is transparent to participants. The initial data launch is occurring 2 years after the national launch of participant enrollment, and includes robust security practices for participant protection. The launch is aligned with the FAIR data principles, providing access to all researchers whose institutions sign a data access agreement (initially targeted to U.S. academic institutions and other health research nonprofits^11^, with plans to include industry and international researchers in the future). For reusability, the cloud-based Jupyter Notebooks platform allows sharing and reproduction of research findings. To encourage reproduction and reuse, the program is making the code of all demonstration analyses presented here available. Additionally, the use of the OMOP Common Data Model, supported by a broad coalition of users, sets a foundation for interoperability of code with other cohorts. By making powerful computational tools available with the data, *All of Us* expands researcher access to those who do not have resources to store and compute on large datasets, and provides a foundation for the future addition of storage-intensive data types such as whole genome sequencing and digital health technologies. To increase transparency of research using the *All of Us* data to the public, including study participants as proscribed in the 21st Century Cures Act, the public Research Hub website includes a description of each Workspace within the Workbench and a directory of all researchers approved for access, both of which are updated every business day (∼24 hours). *All of Us* is also committed to direct return of results to participants, including engagement with participant ambassadors in the dev

Demonstrating responsible curation and research use is also important to earn participants’ trust and show how their data might be used to further health research. The projects described here aim to replicate prior findings and show how the dataset can be used, without preempting novel discoveries. We provide example visualizations of general cohort characteristics to illustrate the heterogeneity of available data types and the diversity of the cohort. Regarding data availability, EHR data are currently available for roughly half of the cohort at this time; this proportion will grow. Response rates to the second set of surveys have increased 37% (from 45,676 to over 62,000) since curation of this dataset and going forward those surveys will be immediately available to participants at enrollment without a 90 day delay. Significant retention efforts are underway including development of a reassessment survey to monitor outcomes over time. Furthermore, nearly 100% of these participants have a biospecimen available and generation of genomic data on this cohort will soon begin. Given the high proportion (over 75%) of participants included from groups traditionally underrepresented in biomedical research, this cohort will provide the foundation for genome-based studies in minority populations and may eventually improve disparities present in genomic medicine today.

The demonstration projects described here, together with the availability of the analysis code for researcher reuse, show the potential of the cohort for a variety of research purposes. For example, the description of medication sequencing in common complex diseases such as T2D and depression speaks to the validity of the data in showing expected treatment patterns, despite gathering data from over 30 individual healthcare facilities. The detailed code provided with this result gives researchers the foundation to extract medications from the data model and extrapolate to classes using a common medication ontology, an approach proven useful in many discovery studies. The PheWAS study shown, while investigating a heavily studied exposure (smoking), shows the power of gathering multiple data sources for a single phenotype, providing researchers with options for study design and validation. While many conclusions about smoking exposure appear mundane now, it is remarkable that these associations can be clearly seen after a few weeks of analyses, when the original discovery took decades. Replicating results is easier than new association discovery, but these studies provide validation validation of the study design that results are consistent with previous findings. Other discoveries may be advanced in this growing dataset, with the entire PheWAS package now available in the Workbench for researcher reuse and new hypothesis generation. Finally, the calculation of the ASCVD pooled risk scores also shows detailed derivation of multiple data elements required for this estimation. While the cohort certainly demonstrates survivor bias in those included, the replication of known race relationships to established risk models and ongoing ability to monitor may provide valuable baseline data for decades to come.

Because participants contribute data in many different ways, the cohort enables prospective, retrospective, and cross-sectional analyses. The COVID-19 crisis occurring concurrently with the planned launch of the Researcher Workbench has provided *All of Us* the opportunity to rapidly adapt and serve the emergent need for COVID-19 relevant research. As the in person enrollment of participants has paused, the consortium has pivoted to use biospecimens for antibody testing to localize early spread of the virus, has developed new surveys that align with other cohorts to gather data directly from participants, and worked to ensure appropriate capture of COVID-19 relevant EHR data. By launching the Researcher Workbench, *All of Us* ensures an appropriate platform will be ready to analyze these critical data in a timely fashion.

These replication projects highlight the value of EHR data from many healthcare partners merged with direct participant data from measurements and surveys and made available in a privacy-sensitive, secure, powerful compute environment. The demonstrations shown here are a small fraction of the extensive tutorials and support created in the Workbench to enable successful discovery research. Workbench support services also include an interactive monitored user forum to communicate with the program and other researchers, as well as an integrated help desk ticketing system to support researchers and gather feedback for the program. Furthermore, the *All of Us* Consortium has completed over 30 additional demonstration projects in additional health focus areas, to further characterize and describe the resource. When published, the analytic code for these projects will also be made directly available to researchers in the Featured Workspaces section of the Workbench for replication and reuse.

Limitations of the beta launch platform presented here include access that is more restricted than initially intended. Finding the balance of privacy, security, and sharing is ongoing, and to fulfill the pledge to protect participants, limiting the beta launch to traditional research groups gives the program an opportunity to learn how to share widely and wisely. Additional risk in this limited launch is exclusion of health disparities and minority health researchers; questions asked during the researcher registration process and Workspace descriptions will provide valuable data to the program regarding diversity of research topics as well as the researcher community. The speed of sharing this dataset just 2 years after launch means that many data types are limited. The survey response rate is low but increasing with earlier delivery of all surveys and focused retention efforts. EHR data missingness is high as is a challenge in many studies, and specific efforts are focused on improving conformance to the data model and completeness from existing sites, as well as newer direct links such as Apple HealthKit and Sync for Science. Additionally, the calculations to balance reidentification risk will be updated as the cohort grows with fewer generalizations expected to be required in future data releases, allowing more granular inspection of groups traditionally underrepresented in research. Finally, as many other large cohorts are developing, including the UK Biobank and the Million Veteran Program, learning to interoperate and jointly analyze data will be paramount. The opportunities to develop novel methodologies to handle data at scale are greater than ever, and the low cost, secure Researcher Workbench platform fulfills a great unmet need to advance precision medicine research.

While significant progress has been done to allow safe sharing of *All of Us* data with the research community, many challenges lie ahead in navigating the future of *All of Us* research, including ensuring ongoing engagement with diverse participants, reduction of data missingness, and rapid expansion of data types including digital health technology, genomics, and external data linkages. The beta launch of the Researcher Workbench begins a process of iterative improvement, fulfilling the goal of providing data to researchers early and often. The *All of Us* Research Program looks forward to incorporating feedback from the research community on this initial release of data and tools.

## Conclusions

The initial release of the *All of Us* Research Program data reflects diverse participants with broad information, reproduces known associations, and provides rich opportunities for research.

The dataset and tools form a strong foundation for cohort growth and future research advancing the program mission to improve human health and advance precision medicine.

## Data Availability

The dataset was accessed through the All of Us Researcher Workbench platform, a cloud-based analytic platform custom built by the program for approved researchers. The Workbench is built on top of the Terra platform, which is also utilized for a number of other NIH-funded studies including the NCI Cloud Resources, the NHLBI BioData Catalyst, and the NHGRI AnVIL. Access to the Researcher Workbench and data are free. Compute and storage accrue usage cost. All researchers who accessed the data for analyses were authorized and approved via a 3-step process that included registration, completion of ethics training, and attestation to a data use agreement. The Researcher Workbench uses Google Compute Engine for computational resources in the cloud and Google Cloud Storage for storage in the cloud.

https://workbench.researchallofus.org/workspaces/aou-rw-dd7cff0e/medicationspathwaysequencesbyracephase1/notebooks

https://workbench.researchallofus.org/workspaces/aou-rw-a8fc912d/duplicateofframinghamahariskscore/notebooks

https://workbench.researchallofus.org/workspaces/aou-rw-d59956e4/jamaphewasfinalreview05212020/data

## Acknowledgments

The *All of Us* Research Program is supported (or funded) by grants through the National Institutes of Health, Office of the Director: Regional Medical Centers: 1 OT2 OD026549; 1 OT2 OD026554; 1 OT2 OD026557; 1 OT2 OD026556; 1 OT2 OD026550; 1 OT2 OD 026552; 1 OT2 OD026553; 1 OT2 OD026548; 1 OT2 OD026551; 1 OT2 OD026555; IAA #: AOD 16037; Federally Qualified Health Centers: HHSN 263201600085U; Data and Research Center: 5 U2C OD023196; Biobank: 1 U24 OD023121; The Participant Center: U24 OD023176; Participant Technology Systems Center: 1 U24 OD023163; Communications and Engagement: 3 OT2 OD023205; 3 OT2 OD023206; and Community Partners: 1 OT2 OD025277; 3 OT2 OD025315; 1 OT2 OD025337; 1 OT2 OD025276. In addition to the funded partners, the All of Us Research Program would not be possible without the contributions made by its participants.

